# A rapid systematic scoping review of the levels of bacterial antimicrobial resistance and antibiotic use amongst people in contact with the criminal justice system

**DOI:** 10.1101/2025.05.15.25327687

**Authors:** Clare Oliver-Williams, Maria Nasim, Michael Cook, Chantal Edge, Diane Ashiru-Oredope

## Abstract

**Background:** Antimicrobial resistance (AMR) poses a significant public health threat. Individuals in contact with the criminal justice system may be particularly vulnerable due to living conditions, behaviours, and pre-existing health issues. This review assesses bacterial AMR and antibiotic use in this population.

**Methods:** A rapid systematic scoping review was conducted (OSF: https://doi.org/10.17605/OSF.IO/XHCFJ). Embase, Medline, and Scopus were searched for studies published between 1 January 2010 and 28 September 2023. One author screened all records, with 10% dual-screened. Included studies examined AMR bacteria or antibiotic use among people in contact with the criminal justice system. Study quality was assessed using the Newcastle-Ottawa Scale and STROBE AMS checklist. Findings were synthesized narratively.

**Results:** Sixteen papers met inclusion criteria; eight were at lower risk of bias. Three studies examined antibiotic use, reporting common inappropriate prescribing (n=1) and associations between recent antibiotic use and resistant infections (n=2). Fourteen papers reported AMR findings, most with a focus on *Mycobacterium tuberculosis* and *Staphylococcus aureus*. Drug-resistant TB prevalence in prison populations ranged from 5.2% to 37% (n=4). MRSA colonization ranged from 8.1% to 8.8% (n=4). Other bacteria examined included *Salmonella spp*., *Acinetobacter spp*., *Group A Streptococcus*, and *Mycoplasma genitalium*.

**Conclusions:** People in contact with the criminal justice system face heightened risks of resistant bacterial infections. However, with only three studies addressing antibiotic use, evidence is limited. Addressing AMR in this group requires collaborative and targeted public health interventions.

## Introduction

Antimicrobial resistance (AMR) presents a serious public health threat^1^. Reasons for the increase in AMR include overuse and misuse of antibiotics in humans and animals, poor infection control practices which allow infections to spread, movement of people, goods, and food across borders which facilitates the global spread of AMR, and a lack of new antibiotic drugs^1,2^.

Individuals in contact with the criminal justice system, defined here as people in custody, prisons, jails or youth offending institutes, may be especially vulnerable to infections^3^. Transmission of organisms, including AMR organisms, may be frequent due to the enclosed, and often overcrowded nature of prisons, the use of communal facilities^3^ and challenges accessing healthcare. The risk of resistant infections may also be increased due to previous or current risk factors such as sex work or injection of illegal drugs prior to entering into the criminal justice system, and also when in the system (e.g. self-harm^4^).

Antibiotic use is a critical factor in the development of AMR. In high-income countries, many correctional facilities have on-site pharmacies. Therefore, exposure to antibiotics in correctional settings may not differ from in the community. Other challenges are present though. Antibiotics may be prescribed with limited diagnostic facilities^5^, and consistent and appropriate treatment may be difficult to achieve, partly because those in prison have limited autonomy to hold their own meds and adhere to dosing schedules which could increase the risk of resistance developing^6^.

People in contact with the criminal justice system are one of several inclusion health groups who experience social exclusion and often have multiple overlapping risk factors for poor health. In addition to people in contact with the criminal justice system, inclusion health groups include: people with drug and/or alcohol dependence; victims of modern slavery; people who sell sex; vulnerable migrants and refugees; Gypsy, Roma, and Traveller communities; and people experiencing homelessness^7^.

Crucially, these groups are not mutually exclusion and individuals may identify with multiple groups at a time point or during their lifetime. This makes it crucial to understand the barriers that not just one but all groups face. This is why this review forms a wider project, seeking to understand AMR, antimicrobial use and antimicrobial stewardship strategies across all inclusion health groups. This review, and the wider project, reflects the UKHSA’s focus on tackling inequalities related to AMR and aligns with the UKHSA’s health equity strategy.

Although, it’s widely suspected that levels of resistant infections may be higher among those inclusion health group, including people in contact with the criminal justice system, to the authors knowledge there are no comprehensive reviews which summarises both the levels of AMR and antibiotic use within these groups. Therefore, we aimed to synthesis all available observational evidence on this topic, to understand the levels of bacterial AMR and antibiotic use among individuals in contact with the criminal justice system.

## Methods

This review was conducted in accordance with the PRISMA guidelines^8^ (**eTable 1**). The protocol for this review is registered on OSF (DOI: https://doi.org/10.17605/OSF.IO/XHCFJ).

**Table 1a:** Summary of studies assessing antimicrobial use among individuals in contact with the criminal justice system.

| Study (country) | Study design | Years of data collection | Source of data | Total *n* | Definition of antibiotic use | Participant characteristics |  |  |  |
| --- | --- | --- | --- | --- | --- | --- | --- | --- | --- |
|  |  |  |  |  |  | Age | Sex | Ethnicity | Length of stay |
| **Szeto et al. 2017 (USA)** | case-control | April 2011 to April 2015 | Medical records | 2020 | Antibiotics use in the past 6 months | mean: 35.9y | Male: 90.3% | Non-Hispanic White 10.0%; Non-Hispanic Black 55.5%; Hispanic 31.4%; Other 3.1% | Median: 126 days |
| **Haysom et al. 2019 (Australia)** | nested case-control | February 2013 to January 2015 | Data collected for the study | 72* | Recent/current antibiotic use: oral antibiotics taken at the time of swab or in the 6 weeks prior to swab. | mean: 16.9y | Male: 92.2% | Indigenous: 59.7%; Non-Indigenous: 40.3% | NG |
| **Di Giuseppe et al. 2021 (Italy)** | case-control | March 2021 to June 2021 | Medical records | 311 | Appropriateness of antibiotic prescription patterns was assessed according to international guidelines ** | mean: 41.8y | Male: 100% | NG | Detained for two years or less: 56.1% |

NG – not given; USA – United States of America
\* Study included 77 discrete SSTI episodes in 72 young people, with 77 instances of recent/current antibiotic use reported in the study
\*\* Centers for Disease Control and Prevention, National Center for Emerging and Zoonotic Infectious Diseases (NCEZID), Division
of Healthcare Quality Promotion (DHQP). Adult Outpatient Treatment Recommendations; Lockhart et al. Evidence-based clinical practice guideline on antibiotic use for the urgent management of pulpal- and periapical-related dental pain and intraoral swelling: A report from the American Dental Association. J. Am. Dent. Assoc. 2019, 150, 906–921; Antimicrobial Stewardship Federal Bureau of Prisons Clinical Guidance

**Table 1b:**
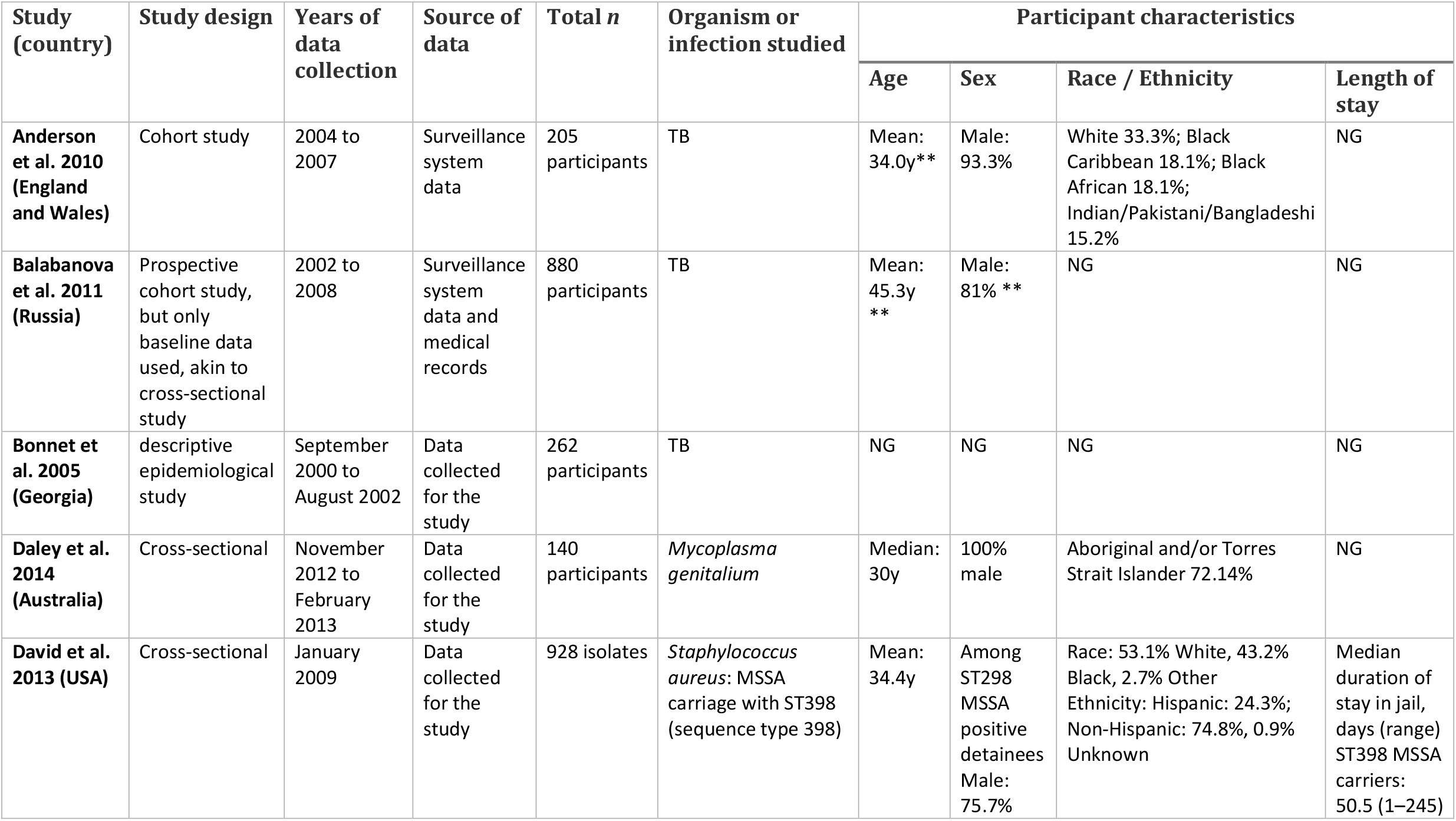
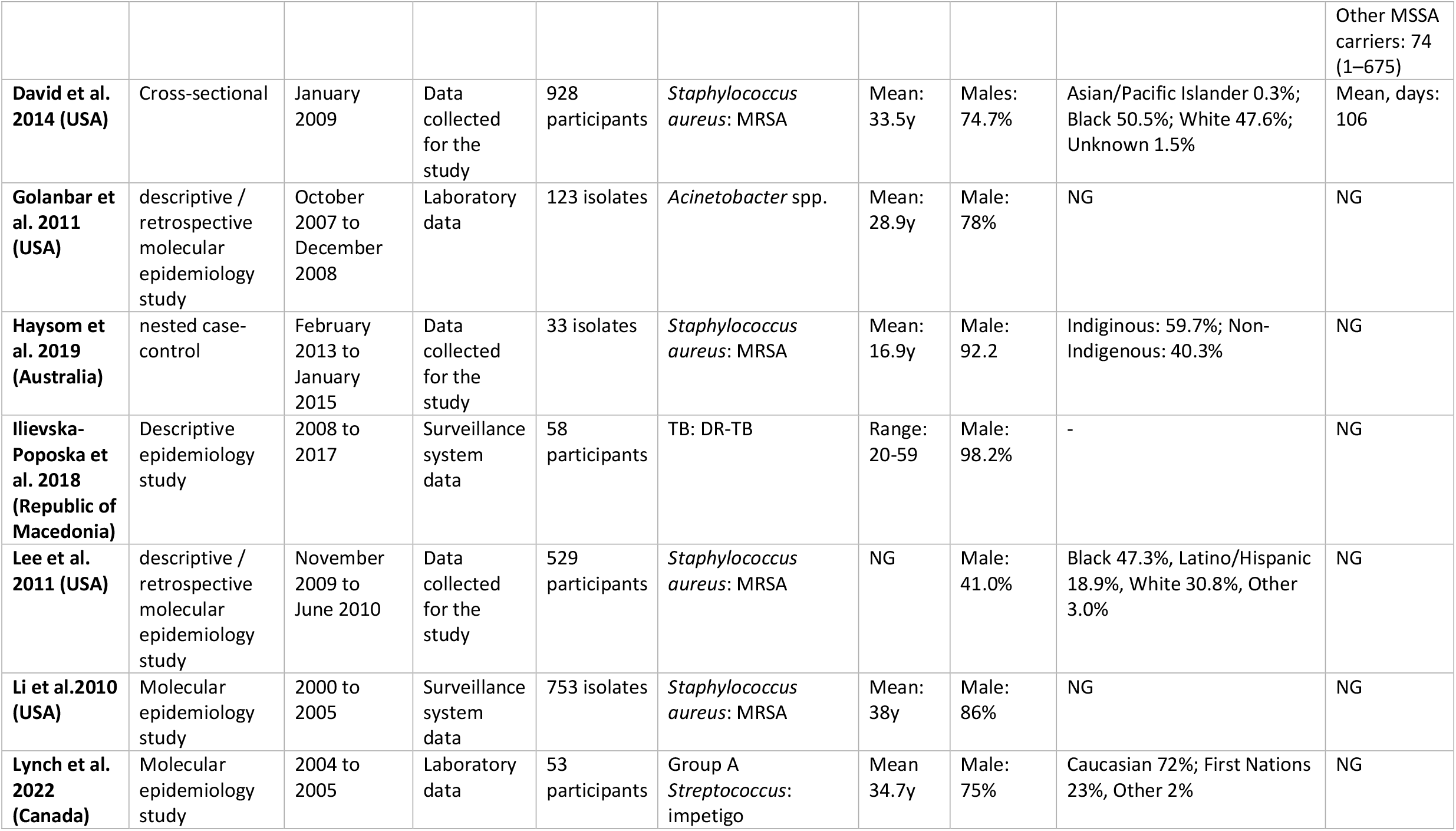
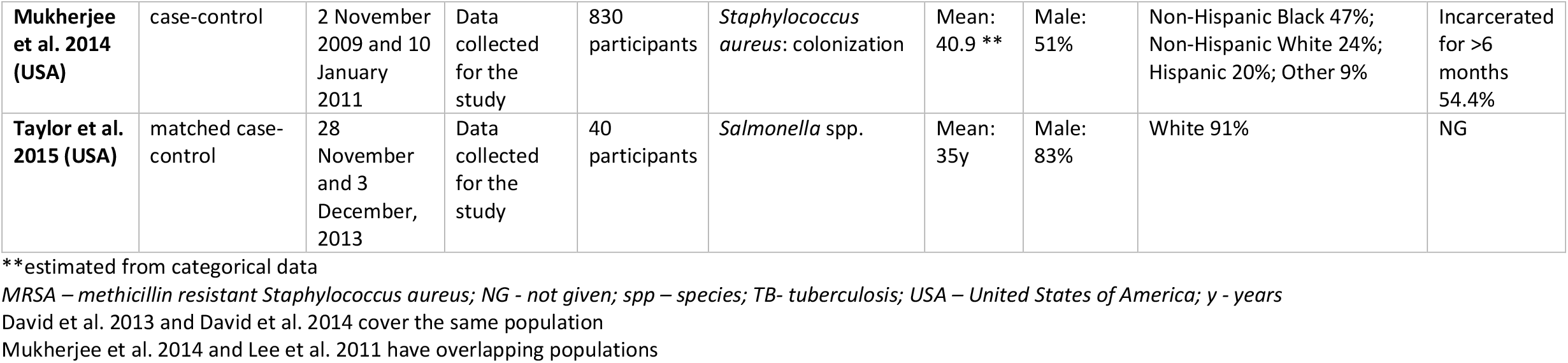
Summary of studies assessing antimicrobial resistant infections among individuals in contact with the criminal justice system.

### Search strategy

The Embase, Medline and Scopus electronic databases were searched for publications between1^st^ January 2010 up to 28^th^ September 2023. The computer-based searches combined terms related to the population (e.g. prisoner*, offender*, jail*) and the outcomes of interest (e.g. Drug Resistance, MDR, antibacterial resist*). Studies could come from any high-income or middle-income country (as set forth by the World Bank^9^), but an English language restriction was placed on the search. Both medical search headings and open text fields were used to identify articles.

Details of the search terms used in Medline are provided in **eTable 2**. As this review comprises part of a larger group of searches, the search terms also include terms for other inclusion health groups. These were screened out prior to initiating this review.

**Table 2:**
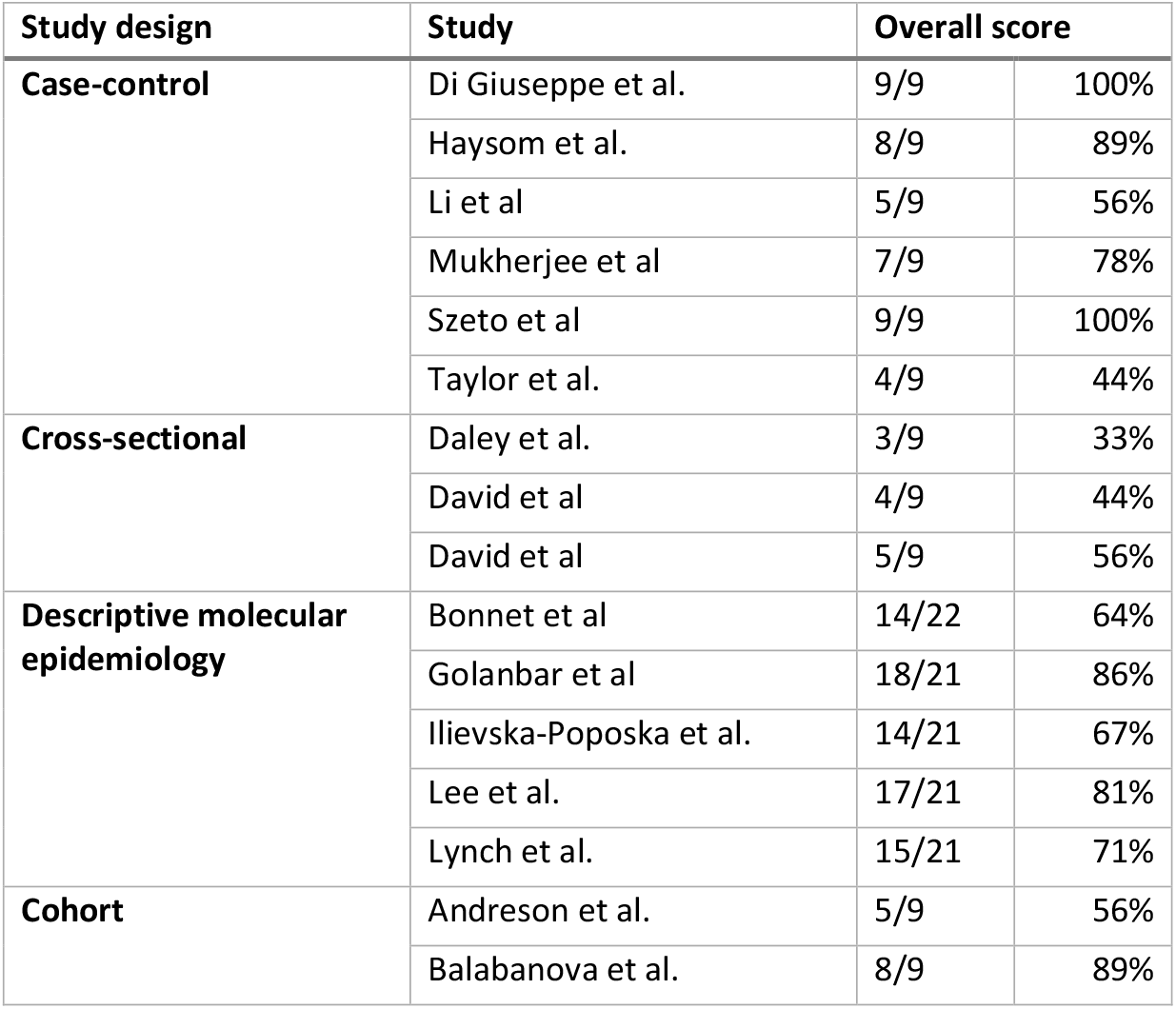
Summary of the results of the study quality assessment of included studies.

### Study selection

To be included in the review, articles had to include either: (1) quantitative information on bacterial AMR among people in contact with the criminal justice system (e.g. the proportion of infections that were resistant), or (2) quantitative information about antibiotic use among people in contact with the criminal justice system (e.g. the percentage of the population who had used antibiotics in the previous year). No age restrictions were applied, as adolescents may be included in the target population. Exclusion criteria were: (1) Case reports, opinion pieces, reviews and editorials as they do not have sufficient or any original data, and (2) studies from low-income countries, as the epidemiology and criminal justice system are more distinct.

All identified abstracts were screened by one author (CO) with 10% of abstracts dual screened by a second author (MN) using the software Rayyan^10^. There were no discrepancies between the authors. For relevant abstracts, full texts were accessed to assess their eligibility for inclusion in the review.

All full text articles were assessed by one author (CO) with dual assessment of 10% by a second author (MN). When disagreements arose in applying the inclusion and exclusion criteria, the authors resolved them through discussion.

### Data extraction

Data was abstracted from the included articles by one author (CO) using a predesign data extraction form. Dual extraction and study quality assessment was performed for a quarter of full text articles by a second author (MN) with any differences discussed and resolved by a third author (XX).

The following data was abstracted: First author and year of publication, year of study, source of data, geographical location of the study, study design, sample size, demographic characteristics of participants, infection or organism studied, case definition, study methodology, outcome studied, study duration and follow up, and results. If data was not available this was recorded in the form. The template data extraction form and the data extracted from included studies are available upon request (**eTable 3**).

Extracted outcome data included the frequency of antibiotic use and bacterial AMR. The latter could include the proportion of infections or organisms that were resistant to specific antibiotics, for example, or the proportion of individuals with an AMR infection. Effect measure extracted depended on the study design, but could include percentages and p-values, odds ratios or risk ratios.

Study quality was assessed using the Newcastle-Ottawa Scale^11^ for cohort, case-control and cross-sectional studies. This is a semi-quantitative scale for assessing the quality of observational studies and allocates a maximum of nine stars to a study, across three categories. The categories are: participant selection criteria, comparability of cases and controls, and exposure assessment (for case-control studies) or outcome assessment (for cohort and cross-sectional studies). The STROBE AMS checklist^12^ was used to assess the quality of outbreak reports and descriptive epidemiology studies, due to a lack of study quality assessment tools tailored to this study design.

### Data synthesis

Due to heterogeneity between the studies with respect to demography, geography and infection/organisms studied, a meta-analysis was deemed inappropriate. Descriptions of the included studies were tabulated, and a narrative synthesis was performed. Findings relating to bacterial AMR and antibiotic use were summarised separately. Within AMR infections, studies assessing the same infection or organism were summarised together.

## Results

The search identified 521 papers; 431 were excluded during abstract screening. The remaining 90 papers were reviewed in full. Sixteen papers were included in the final review. (**Figure 1**). **Table 1a** summarises the included antibiotic use studies and **Table 1b** summarises the included bacterial AMR studies. Two cohort studies^13,14^, three cross-sectional studies^15–17^, five molecular epidemiology studies^18–22^ and six case-control studies^23–28^ were identified.

**Figure 1:**
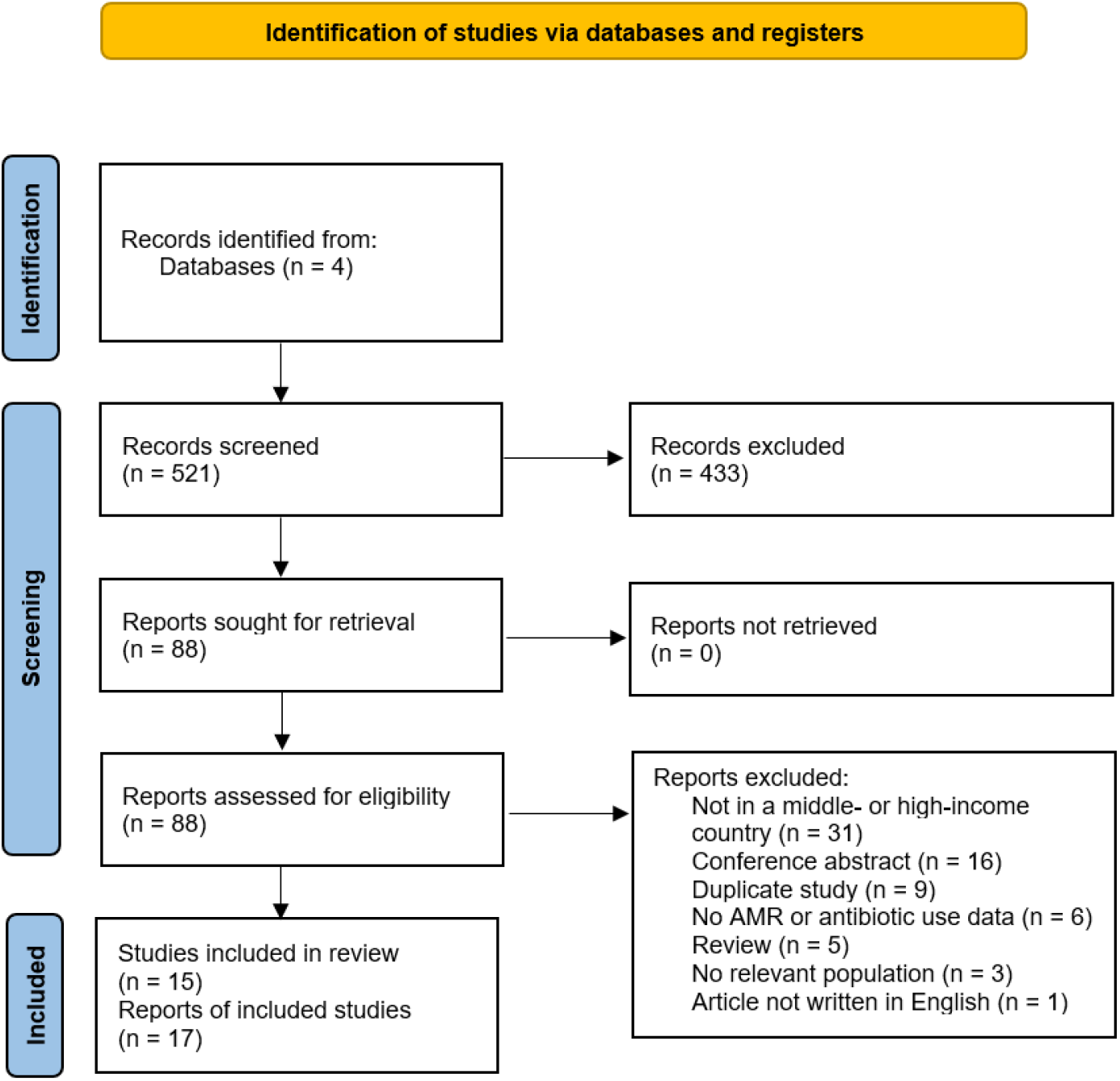
Prisma flow diagram summarising the identification of relevant studies AMR – Antimicrobial Resistance

Three papers reported findings relating to antibiotic use^24,27,28^, and 14 papers reported AMR results^13–23,25–27^, covering *Acinetobacter spp*. (1 paper^19^), Group A *Streptococcus* (1 paper^22^) *Mycoplasma genitalium* (1 paper^17^), Salmonella spp. (1 paper^23^), tuberculosis (4 papers^13,14,18,21^), and *Staphylococcus aureus* (6 papers^15,16,20,25–27^).

The findings of the study quality assessment can be found in **Table 2**, with detailed breakdown of scores in **eTables 4-6**. One cohort study (Balabanova et al.^14^) was of high study quality scoring 89%, but the second cohort study (Andreson et al.^13^) did not score as highly (56%) indicating a greater risk of study bias. None of the three cross-sectional studies (David et al, David et al. and Daley et al.^15–17^) scored highly, and are more likely to be biased. Two case-control studies (Taylor et al. and Li et al.^23,26^) did not score highly, but the remaining four studies did (Di Giuseppe et al., Haysom et al., Mukherjee et al. and Szeto et al.^24,25,27,28^). Of the five molecular epidemiology studies, three studies scored above 70% for study quality (Lee et al., Golanbar et al., and Lynch et al.^19,20,22^] with the final two studies (Ilievska-Poposka et al. and Bonnet et al. ^18,21^) scoring 67% and 64%, respectively.

## Antibiotic use studies

Three studies were identified that assessed antibiotic use among people in contact with the criminal justice system. Two case-control studies were identified, and one nested case-control study.

Szeto et al.^24^ included 2020 individuals in a study based in the New York City jail system. Of the 1010 individuals with a skin and soft tissue infection (SSTI), 28% had used antibiotics in the previous 6 months. The percentage was lower among the 1010 individuals without a skin and soft tissue infection (11.1%). Overall, 19.6% of participants used antibiotics in the previous 6 months.

Di Giuseppe et al.^28^ assessed 311 prisoners in the South of Italy for the appropriateness of prescribing for upper respiratory tract infections or dental infections. In 77.5% of diagnoses an antibiotic was prescribed, with mean duration of 5.4 days (SD ± 1). In 69.4% of cases where an antibiotic was not clinically indicated antibiotics were prescribed, whereas an antibiotic prescription was provided in all 52 cases in which it was indicated. The most frequently prescribed antibiotic was amoxicillin with clavulanic acid (33.8%), followed by amoxicillin alone (26.5%), macrolides (19.8%) and third generation cephalosporins (7.9%).

Haysom et al.^27^ included 72 young people in a Juvenile Custodial Centres, who collectively had 77 discrete episodes of SSTI, such as boils, abscesses, impetigo, cellulitis or surgical wound infections. Thirty-three isolates tested positive for MRSA. Those with MRSA SSTI were significantly more likely to have used oral antibiotics taken at the time of swab or in the 6 weeks prior to the swab (36.4%) compared to those with non-MRSA SSTI (13.6%; p<0.05).

### Bacterial AMR studies

#### Tuberculosis

Four studies assessing AMR TB infections were identified^13,14,18,21^; two molecular epidemiology studies^18,21^ and two cohort studies^13,14^.

Bonnet et al.^18^ collected data and sputum samples from 262 participants in Abkhasia prison in Georgia. Of the 262, 122 were new TB cases, 140 were previously treated cases. 35.2% of new cases and 57.6% of previously treated cases had any kind of resistance.

The proportion of individuals with TB resistant to isoniazid was 37%. Among the 122 new cases, 23.8% of cases had some resistance (mono, poly or multi-drug resistance) to isoniazid, and 5.7% had resistance to rifampicin. 4% of new cases were MDR. For the 140 previously treated cases, 47.9% had any resistance to isoniazid and 20% had any resistance to rifampicin. 18.7% of previously treated cases were MDR.reported.

Ilievska-Poposka et al.^21^ identified 58 TB cases across all prisons in the Republic of Macedonia. Of the 58 cases, 5.2% had DR-TB. One was resistant to Ethambutol (1.7%), one resistant to Streptomycin and Isoniazid (1.7%) and one was resistant to Streptomycin, Ethambutol and Rifampicin (1.7%).

There were no cases with MDR-TB

Balabanova et al.^14^ assessed susceptibility to first-line TB drugs for 67 isolates from prisoners in Russia. Approximately half of cases had MDR-TB (49.2%)

Anderson et al.^13^ used TB surveillance data which covered all prisons in England and Wales. Of the 136 cases tested, 1 had MDR (7.6%), 34.5% were isoniazid resistant.

#### Staphylococcus aureus

Six studies assessing AMR *S. aureus* infections were identified^15,16,20,25–27^; two cross-sectional^15,16^, one molecular epidemiology study^20^ and three case-control studies^25–27^.

David et al. published two papers assessing the MRSA carriage of individuals in Dallas County Jail, USA. David et al. 2013^15^ looked at 20ST398 MSSA isolates from 16 detainees, 18 out of 20 isolates were susceptible to ciprofloxacin, gentamicin, rifampin, and trimethoprim-sulfamethoxazole and resistant to clindamycin and erythromycin. The remaining two isolates differed: one was susceptible to clindamycin and the other was resistant to ciprofloxacin.

David et al. 2014^16^ assessed hand and nasal *S. aureus* carriage among 928 individuals. 8.1% of individuals had MRSA hand and/or nasal carriage (6.3% nasal carriage, 4.1% hand carriage). The proportion of individuals with MSSA carriage was much higher: 34.3% had MSSA hand and/or nasal carriage (26.5% nasal carriage, 20.7% hand carriage). 58.7% of individuals were *S. aureus* negative

Among subjects with nasal MRSA carriage, 41% had hand MRSA carriage, and 29% of those with hand MRSA carriage did not have nasal *S. aureus* carriage.

When MRSA clustering within the 68 detention divisions (known as tanks) was assessed, the prevalence of MRSA colonization ranged from 6.4% to 50% in the 42 tanks where MRSA was identified. In most of the tanks (74%) detainees did not carry genetically concordant MRSA strains. In the other 26% of tanks, concordance ranged from 8-37%. Thus, there was limited evidence of frequent spread of MRSA among detainees Of the demographic factors assessed, MRSA carriage only varied by race, with a higher proportion among Black individuals compared to White individuals (10.5% vs 5.7%, p=0.008; odds ratio=2.7; 95% confidence interval [CI], 1.5-5.0; p=0.002)). No difference was found in length of stay, age or gender. Carriage of an *S. aureus* isolate was not associated with age or length of jail stay, but black females were about half as likely as white males to be colonized with *S. aureus* (odds ratio=0.51; 95% CI, 0.33-0.79; p=0.003).

Mukherjee et al.^25^ conducted a case-control study assessing nasal and oropharyngeal colonization by *S. aureus* in 830 inmates entering two maximum-security prisons in New York State, USA. The prevalence of nasal colonization with MRSA was similar for both females and males (2.7% and 2.4%, respectively), and little difference was found when looking at oropharyngeal colonization either (3.5% vs 2.6%). However, larger differences in the colonization of both nasal and oropharynx was between females (4.4%) and males (0.9%). Thus, 10.6% of females and 5.9% of males (8.2% overall) were colonized at either or both sites.

Multivariable regression analysis did not identify any risk factors for MRSA colonization and risk characteristics in women or men entering prison (factors assessed were age, self-reported health, incarceration duration, shared personal items, skin infection within past 6 months and social group membership).

Lee et al.^20^ also looked at MRSA colonization within the same population. They found that 30% of individuals who tested *S. aureus* positive in both the nasal and oropharyngeal samples had different MRSA strains (identified by their spa types). Thus, they were classified as dually colonized. This included 10 individuals colonized with both MSSA and MRSA.

Li et al.^26^ conducted a molecular epidemiology study, looking at 753 isolates identified through surveillance of the four state-managed community correctional centers (jails) and four state-managed correctional facilities in Hawai’i, USA.

A steady increase in the proportion of *S. aureus* isolates that were MRSA was observed for the total inmate population during the 6-year study period, from 48% in 2000 to 74% in 2005 (p<0.0001), which was found among individuals in jail (from 50.0% to 75.3%; p<0.001), and those in prison, although the latter was not significant (from 46.9% to 70.5%; p=0.0549). The proportion of MRSA resistance to clindamycin decreased over the time period, but an increase in the proportion of MRSA resistant to erythromycin was found (p<0.0001). MRSA resistance to other antimicrobials was generally either low or non-exist. No significant difference was observed in the MRSA proportion between male (69%) and female (70%) inmates.

A significant increase in the incidence of MRSA was observed among jail inmates between 2000-2005: 3.2/1000 to 88.4/1000 (p=0.005). This is an annual average increase of 106%. A statistically non-significant increase was observed among prison inmates from 7/1000 to 18.3/1000 during this same period (p=0.18).

Haysom et al.^27^ collected 33 MRSA isolates from SSTI from individuals in Juvenile Custodial Centres. All isolates were sensitive to co-trimoxasole and vancomycin and 91% were sensitive to clindamycin.

#### Other infections

Findings for Group A *Streptococcus, Acinetobacter spp*., *Mycoplasma genitalium* and *Salmonella spp*. were reported in separate papers^17,19,22,23^.

Lynch et al^22^ collected isolates from impetigo lesions in 53 adults and tested for AMR genes (*aph (3’)-III*; *erm* (A); *tet* (M)). Three AMR genes were identified across 9 of the 53 isolates (17,0%), all genes belonged to genomes in the *emm* type groups 114 (8/53 isolates) and 114.1 (1/53 isolate).

Taylor et al.^23^ tested faecal samples from 40 participants for Salmonella strains. Nine tested positive (22.5%). Of the nine, two isolates displayed resistance to five drug classes, including third generation cephalosporins.

Golanbar et al.^19^ assessed 123 isolates that tested positive for *Acinetobacter spp*. across all California Correctional Facilities. The five least effective antibiotics were ceftriaxone (71%), cefotaxime (64%), piperacillin (48%), tetracycline (36%), and ceftazidime (36%). Antibiotics with high potency against most of the isolates were polymyxin B (100%), tigecycline (94%), minocycline (93%), amikacin (90%), and meropenem (89%). Multidrug-resistant isolates were found in only six facilities. Furthermore, the presence of class 1 integrons was detected in 15 isolates, all of which are MDR.

Daley et al^17^ tested urine samples from 140 men for *Mycoplasma genitalium*. Eight out of 140 [5.71% (95% CI 1.82–9.60)] samples tested positive, and two out of eight (25%) samples carried a mutation in the 23S rRNA gene associated with macrolide resistance.

## Discussion

AMR has emerged as a global public health threat, posing significant challenges to the effective treatment of infectious diseases^1^. While extensive research has been conducted on AMR in hospitals and communities, this review aimed to address the gap in our understanding of the prevalence of AMR infections and antibiotic among those in contact with the criminal justice system and any disparities The review highlights that the levels of AMR infections vary by country (from 5.2% of TB infections in Macedonia^21^ to 37% in Russia^18^). These differences may reflect country prevalence or inter-country differences in risk factors prior to or in prisons, as well as differences in study design and sampling.

No studies compared AMR levels with those in the general population. However comparisons with previous literature provide some indication of higher risk of AMR infections. For example, rates of MDR-TB (ranging from 7.6% to 49.2%) are higher than in the general population. Globally in 2023, estimates of MDR-TB for new TB cases were 3.2% (95% UI: 2.5–3.8%) and 16% (95% UI: 9.0–24%) in previously treated cases%) in most of the identified studies^29^. estimates of MRSA carriage were more consistent (at approximately 8%) between different forms of custodial settings^15,16,25,26^, which is considerably higher than estimates in the general population^30^.

In one of the three studies of antibiotic use^28^, high levels of over-prescribing were noted. In conjunction with this, one study noted that those with MRSA SSTI were significantly more likely to have taken oral antibiotics in the 6 weeks prior to swabbing compared to those with non-MRSA SSTI^27^, emphasizing the importance of appropriate prescribing practices. This indicates that those in contact with the criminal justice system should be a priority group when tackling AMR, due to potentially high levels of AMR infections, inappropriate prescribing and challenges implementing infection prevention and control programs in correctional settings^31^.

Several factors may contribute to an increased risk of AMR infections among those in contact with the criminal justice system. This includes overcrowded living conditions^32^, challenges in accessing healthcare^6^ and high prevalence of risk factors such as tattooing^33^ and substance abuse^34^.

Furthermore, the frequent movement of inmates between facilities and the community will contribute to the spread of resistant strains. Finally, the prison population have worse health than the general population in multiple ways, including high levels of long term physical and mental illness, and bloodborne virus infections^35^, which complicate the management of AMR infections. Collectively, this underscores the need for comprehensive prevention and treatment strategies tailored to the unique needs of the prison population

As previously noted, people in contact with the criminal justice system are one of several inclusion health groups, and may identify with other inclusion health groups also. Access to quality healthcare is crucial to overcoming the health inequalities noted in this review. Guidance exists to support this. The National Health Service (NHS) framework on inclusion health sets out how to provide quality healthcare for all, with an emphasis on equitable access, positive healthcare experiences and optimal outcomes^7^. The Homeless and Inclusion Health standards for commissioners and service providers proposes minimum standards for planning, commissioning and providing healthcare for excluded groups^2^.

This review has a number of strengths. This includes a detailed, comprehensive search strategy, covering three databases, which was inclusive of individuals from all parts of the criminal justice system and all bacterial infections. However, there are also limitations. Firstly, data heterogeneity prevented us from performing meta-analyses to provide an overview of the levels of AMR infections. Second, not all studies were of a high quality. Eight studies were at higher risk of bias. Finally, the limited number of different infections under assessment means that conclusions cannot be drawn about all bacterial infections.

This review has also identified several gaps within the literature. In addition to the limited number of organisms that were assessed (6 across all included papers), there is a paucity of research focusing on individuals in youth offender institutes, with only a single paper identified. Additionally, no studies assessed individuals on probation. These are notable gaps that need to be rectified to better understand these populations. In addition, only one study provided trend data over time, and only three studies provided information about antibiotic use. This limits our ability to understand long-term patterns and identify ways to improve prescribing practices. Finally, it was noted that no papers compared levels of AMR among those in custodial settings with those in the general population. Without direct comparison it is more challenging to understand the magnitude of the inequalities experienced by people in contact with the criminal justice system.

Despite this, the high rates of bacterial AMR found in this review also have implications for both public health policy and practice. AMR infections within prisons is a complex, multifaceted challenge. It requires multi-agency, collaborative intervention to tackle it. This should include surveillance, prevention, and treatment approaches. Strengthened infection prevention and control measures, strong antimicrobial stewardship programmes tailored to the health and justice setting, improved facilities to support residents to manage their own medication and improved access to healthcare services are all needed to mitigate the spread of resistant bacteria. Long-term collaboration between the prison sector, the healthcare system and public health organisations is needed to support this work, along with training of and awareness raising among residents about the importance of AMR and infection prevention.

In conclusion, individuals in contact with the criminal justice system are a marginalised population at risk of AMR infections. Comparison with the wider literature indicates this risk is higher than in the general population. To tackle this, targeted approaches tailored to the sector are needed to prevent and control their spread amongst the prison population. It is essential to address the unique risk factors and barriers to care that are faced by the prison population, which will require on-going collaboration with public health organisation and the healthcare system. To ensure effective interventions are implemented, continued research and surveillance is also needed to monitor trends and inform policymaking.

## Supporting information

eTable

## Data Availability

All data produced in the present work are contained in the manuscript, and are also available from the original research articles referenced within the review.

