## Supplementary material for "A rapid systematic scoping review of the levels of bacterial antimicrobial resistance and antibiotic use amongst people in contact with the criminal justice system": eTable

| Section and Topic | Item # | Checklist item | Location where item is reported |
|---|---|---|---|
| **TITLE** | | | |
| Title | 1 | Identify the report as a systematic review. | 1 |
| **ABSTRACT** | | | |
| Abstract | 2 | See the PRISMA 2020 for Abstracts checklist. | 2 |
| **INTRODUCTION** | | | |
| Rationale | 3 | Describe the rationale for the review in the context of existing knowledge. | 3 |
| Objectives | 4 | Provide an explicit statement of the objective(s) or question(s) the review addresses. | 3 |
| **METHODS** | | | |
| Eligibility criteria | 5 | Specify the inclusion and exclusion criteria for the review and how studies were grouped for the syntheses. | 4 |
| Information sources | 6 | Specify all databases, registers, websites, organisations, reference lists and other sources searched or consulted to identify studies. Specify the date when each source was last searched or consulted. | 4 |
| Search strategy | 7 | Present the full search strategies for all databases, registers and websites, including any filters and limits used. | Supplementary material, eTable 2 |
| Selection process | 8 | Specify the methods used to decide whether a study met the inclusion criteria of the review, including how many reviewers screened each record and each report retrieved, whether they worked independently, and if applicable, details of automation tools used in the process. | 4 |
| Data collection process | 9 | Specify the methods used to collect data from reports, including how many reviewers collected data from each report, whether they worked independently, any processes for obtaining or confirming data from study investigators, and if applicable, details of automation tools used in the process. | 4-5 |
| Data items | 10a | List and define all outcomes for which data were sought. Specify whether all results that were compatible with each outcome domain in each study were sought (e.g. for all measures, time points, analyses), and if not, the methods used to decide which results to collect. | 4 |
| | 10b | List and define all other variables for which data were sought (e.g. participant and intervention characteristics, funding sources). Describe any assumptions made about any missing or unclear information. | 4 |
| Study risk of bias assessment | 11 | Specify the methods used to assess risk of bias in the included studies, including details of the tool(s) used, how many reviewers assessed each study and whether they worked independently, and if applicable, details of automation tools used in the process. | 4 |
| Effect measures | 12 | Specify for each outcome the effect measure(s) (e.g. risk ratio, mean difference) used in the synthesis or presentation of results. | 4 |
| Synthesis | 13a | Describe the processes used to decide which studies were eligible for each synthesis (e.g. tabulating the study intervention characteristics | n/a |

| Section and Topic | Item # | Checklist item | Location where item is reported |
|---|---|---|---|
| methods | | and comparing against the planned groups for each synthesis (item #5)). | |
| | 13b | Describe any methods required to prepare the data for presentation or synthesis, such as handling of missing summary statistics, or data conversions. | n/a |
| | 13c | Describe any methods used to tabulate or visually display results of individual studies and syntheses. | 5 |
| | 13d | Describe any methods used to synthesize results and provide a rationale for the choice(s). If meta-analysis was performed, describe the model(s), method(s) to identify the presence and extent of statistical heterogeneity, and software package(s) used. | 5 |
| | 13e | Describe any methods used to explore possible causes of heterogeneity among study results (e.g. subgroup analysis, meta-regression). | n/a – too few studies |
| | 13f | Describe any sensitivity analyses conducted to assess robustness of the synthesized results. | n/a |
| Reporting bias assessment | 14 | Describe any methods used to assess risk of bias due to missing results in a synthesis (arising from reporting biases). | n/a |
| Certainty assessment | 15 | Describe any methods used to assess certainty (or confidence) in the body of evidence for an outcome. | n/a |
| **RESULTS** | | | |
| Study selection | 16a | Describe the results of the search and selection process, from the number of records identified in the search to the number of studies included in the review, ideally using a flow diagram. | 6 |
| | 16b | Cite studies that might appear to meet the inclusion criteria, but which were excluded, and explain why they were excluded. | n/a |
| Study characteristics | 17 | Cite each included study and present its characteristics. | 6, Table 1a and 1b |
| Risk of bias in studies | 18 | Present assessments of risk of bias for each included study. | Table 2, eTable 4-6 |
| Results of individual studies | 19 | For all outcomes, present, for each study: (a) summary statistics for each group (where appropriate) and (b) an effect estimate and its precision (e.g. confidence/credible interval), ideally using structured tables or plots. | 6-9 |
| Results of syntheses | 20a | For each synthesis, briefly summarise the characteristics and risk of bias among contributing studies. | n/a |
| | 20b | Present results of all statistical syntheses conducted. If meta-analysis was done, present for each the summary estimate and its precision (e.g. confidence/credible interval) and measures of statistical heterogeneity. If comparing groups, describe the direction of the effect. | n/a |
| | 20c | Present results of all investigations of possible causes of heterogeneity among study results. | n/a – too few studies |
| | 20d | Present results of all sensitivity analyses conducted to assess the robustness of the synthesized results. | n/a |
| Reporting biases | 21 | Present assessments of risk of bias due to missing results (arising from reporting biases) for each synthesis assessed. | n/a |
| Certainty of | 22 | Present assessments of certainty (or confidence) in the body of evidence for each outcome assessed. | n/a |

| Section and Topic | Item # | Checklist item | Location where item is reported |
|---|---|---|---|
| evidence | | | |
| **DISCUSSION** | | | |
| Discussion | 23a | Provide a general interpretation of the results in the context of other evidence. | 10 |
| | 23b | Discuss any limitations of the evidence included in the review. | 10 |
| | 23c | Discuss any limitations of the review processes used. | 10 |
| | 23d | Discuss implications of the results for practice, policy, and future research. | 10-11 |
| **OTHER INFORMATION** | | | |
| Registration and protocol | 24a | Provide registration information for the review, including register name and registration number, or state that the review was not registered. | 4 |
| | 24b | Indicate where the review protocol can be accessed, or state that a protocol was not prepared. | 4 |
| | 24c | Describe and explain any amendments to information provided at registration or in the protocol. | n/a |
| Support | 25 | Describe sources of financial or non-financial support for the review, and the role of the funders or sponsors in the review. | 17 |
| Competing interests | 26 | Declare any competing interests of review authors. | 17 |
| Availability of data, code and other materials | 27 | Report which of the following are publicly available and where they can be found: template data collection forms; data extracted from included studies; data used for all analyses; analytic code; any other materials used in the review. | 5 |

**eTable 1**: PRISMA checklist

1 exp Drug Resistance, Bacterial/ or exp antibiotic resistance/ or Gene Transfer, Horizontal/ or exp Anti-Bacterial Agents/ (892163)

10 or/7-9 (57546)

11 6 and 10 (431)

2 ((antibacterial or antimicrobial or antibiotic or drug or multidrug) adj2 resist*).tw. (245206)

3 (ABR or AMR or MDR or "resistance gene*" or "horizontal gene transfer" or "resistance determinant*" or "multi-resistance" or multiresistance or "antibiotic susceptibility" or resistome*).tw. (97343)

4 (Antibiotic* or prescribing or prescription* or regimen*).ti. or (Antibiotic* or prescribing or prescription* or regimen*).ab. /freq=2 (393631)

5 or/1-4 (1258463)

6 limit 5 to (english language and yr="2010 -Current") (550173)

7 *Prisoners/ or exp *Correctional Facilities/ (20073)

8 (Prison* or imprison* or penitentiar* or jail* or remand or custod* or gaol or detention or penal or criminal* or felon* or incarcer* or detaine* or inmate*).ti. or (Prison* or imprison* or penitentiar* or jail* or remand or custod* or gaol or detention or penal or criminal* or felon* or incarcer* or detaine* or inmate*).ab. /freq=2 (41217)

9 ("correctional facilit*" or "secure estate" or IRC or "immigration removal centre" or convict or convicts or convicted or offender).tw,kf. (5874)

**eTable 2**: Search terms used in Medline

| Lead Author | Year of Publication | Year of Study | Source of data | Location | Study Design (RCT, cohort study, etc) | Sample Size | Population Characteristics (demographics of the study participants | | | | infection, definition of case | Methodology | Outcome | Duration (Study period and the follow up) | Findings | Comments |
|---|---|---|---|---|---|---|---|---|---|---|---|---|---|---|---|---|
| | | | | | | | age | sex | Race/ethnicity | other | | | | | | |

**eTable 3**: Example data extraction form

| Study | Selection | Comparability | Exposure | Findings | |
|---|---|---|---|---|---|
| **Szeto et al** | 4 | 2 | 3 | 9/9 | 100% |
| **Haysom et al.** | 3 | 2 | 3 | 8/9 | 89% |
| **Taylor et al.** | 3 | 0 | 1 | 4/9 | 44% |
| **Di Giuseppe et al.** | 4 | 2 | 3 | 9/9 | 100% |
| **Mukherjee et al** | 3 | 2 | 2 | 7/9 | 78% |
| **Li et al** | 3 | 0 | 2 | 5/9 | 56% |

**eTable 4**: Results of the study quality assessment of included case-control studies.

| Study | Selection | Comparability | Outcome | Overall score | |
|---|---|---|---|---|---|
| **David et al** | 2 | 0 | 2 | 4/9 | 44% |
| **David et al** | 0 | 2 | 3 | 5/9 | 56% |
| **Daley et al.** | 1 | 0 | 2 | 3/9 | 33% |

**eTable 5**: Results of the study quality assessment of included cross-sectional studies.

| Study | Selection | Comparability | Outcome | Overall score | |
|---|---|---|---|---|---|
| **Balabanova et al** | 4 | 2 | 2 | 8/9 | 89% |
| **Andreson et al.** | 4 | 0 | 1 | 5/9 | 56% |

**eTable 6**: Results of the study quality assessment of included cohort studies.

| | Lynch et al. | Golanbar et al. | Lee et al. | Ilievska-Poposka et al. | Bonnet et al. |
|---|---|---|---|---|---|
| **1a. Indicate the study design with a commonly used term in the title or the abstract. Title and Abstract** | Yes | Yes | No | No | No |
| **1b. Provide an informative and balanced summary in the abstract. Title and Abstract** | Yes | Yes | No (research letter) | Yes | Yes |
| **2. Explain the scientific background and rationale for the investigation being reported. Introduction** | Yes | Yes | Yes | Yes | Yes |
| **3. State specific objectives, including any prespecified hypotheses. Introduction** | Yes | Yes | Yes | Yes | Yes |
| **4. Present key elements of study design early in the paper. Methods** | Yes | Yes | Yes | Yes | Yes |
| **5. Describe the setting, locations and relevant dates. Results** | Yes | Yes | Yes | Yes | Yes |
| **6. Description of participants—eligibility criteria, sources and methods. Results** | Yes | Yes | Yes | Yes | No |
| **7. Clearly define all outcomes, exposures, predictors, potential confounders, and effect modifiers. Methods** | No | Yes | Yes | No | Yes |
| **8. For each variable of interest, give sources of data and details of methods of assessment. Methods** | No | Yes | Yes | No | Yes |
| **9. Describe any efforts to address potential sources of bias. Methods** | No | No | Yes | No | No |
| **10. Explain how the study size was arrived at. Methods** | Yes | Yes | Yes | Yes | No |
| **11. Explain how quantitative variables were handled. Methods** | No | Yes | Yes | No | Yes |
| **12. Describe all statistical methods, including those used to control for confounding. Explain how missing data, lost to follow up and sensitivity analyses were addressed. Methods** | No | Yes | Yes | No | Yes |
| **13. Report the numbers of individuals at each stage and reason for non-participation. Results** | Yes | Yes | Yes | Yes | No |

| | | | | | |
|---|---|---|---|---|---|
| **14. Give the characteristics of study participants. results** | Yes | Yes | Yes | Yes | No |
| **15. Report outcome data. Results** | Yes | Yes | Yes | Yes | Yes |
| **16. Give unadjusted estimates and, if applicable, confounder-adjusted estimates. Make clear which counfounders were adjusted for and why they were included. Results** | Yes | Yes | Yes | Yes | Yes |
| **17. Report other analyses performed—eg, analyses of subgroups Results** | n/a | n/a | n/a | n/a | Yes |
| **18. Summarise key results with reference to study objectives. Discussion** | Yes | Yes | Yes | Yes | Yes |
| **19. Discuss limitations of the study, taking into account sources of potential bias or imprecision. Discussion** | No | No | No | No | Yes |
| **20. Give a cautious overall interpretation of results. Discussion** | Yes | Yes | Yes | Yes | Yes |
| **21. Discuss the generalisability. Discussion** | No | No | No | No | No |
| **22. Give the source of funding. Funding** | Yes | Yes | Yes | Yes | No |
| **Overall score** | 15/21 (71%) | 18/21 (86%) | 17/21 (81%) | 14/21 (67%) | 14/22 (64%) |

**eTable 7**: Results of the study quality assessment of molecular epidemiology studies.
